# MIND diet associated with later onset of Parkinson’s disease

**DOI:** 10.1101/2020.07.13.20151977

**Authors:** A Metcalfe-Roach, AC Yu, E Golz, K Sundvick, MS Cirstea, D Kliger, LH Foulger, M Mackenzie, BB Finlay, S Appel-Cresswell

## Abstract

**Background:** The MIND diet has been linked with prevention of Alzheimer’s disease and cognitive decline but has not been fully assessed in the context of Parkinson’s disease (PD).

**Objective:** To determine whether MIND diet adherence is associated with the age of Parkinson’s disease onset in a manner superior to that of the Mediterranean diet.

**Methods:** Food Frequency Questionnaires from 167 participants with PD and 119 controls were scored for MIND and two versions of Mediterranean diet adherence. Scores were compared between sex and disease subgroups, and PD diet adherence was correlated with age of onset using univariate and multivariate linear models.

**Results:** The female subgroup adhered more closely to the MIND diet than the males, and diet scores were not modified by disease status. Later age of onset correlated most strongly with MIND diet adherence in the female subgroup, corresponding to differences of up to 17.4 years (p<0.001) between low and high dietary tertiles. Greek Mediterranean adherence was also significantly associated with later PD onset across all models (p=0.05-0.03). Conversely, only Greek Mediterranean adherence remained correlated with later onset across all models in men, with differences of up to 8.4 years (p=0.002).

**Conclusions:** This cross-sectional study finds a strong correlation of age of onset of PD with dietary habits, suggesting that nutritional strategies may be an effective tool to delay PD onset. Further studies may help to elucidate potential nutrition-related sex-specific pathophysiological mechanisms and differential prevalence rates in PD.

## INTRODUCTION

Numerous epidemiological studies have investigated the effects of regional dietary trends on population health and longevity. The Western diet, common in North America, is notorious for its high levels of processed and fried foods, sugar, and red meat; this diet has been linked to increased prevalence and severity of many diseases, including cardiovascular disease (CVD), diabetes, and cancer^1,2^. Conversely, the Mediterranean diet (MeDi) has garnered significant interest due to its association with reduced rates of cancer^3^, CVD^3^, and neurodegenerative diseases^4^ such as Alzheimer’s disease (AD) and Parkinson’s disease (PD). Two principal MeDi scoring methods exist: the original MeDi (OMeDi) is characterized in part by its antioxidant-rich mix of vegetables, whole grains, and reduced red meat/dairy^5^ and was revised to promote fish intake, while the alternative Greek MeDi (GMeDi) pattern uses similar food groups but also promotes potato intake and limits poultry consumption^6^.

The Mediterranean-DASH Intervention for Neurodegenerative Delay (MIND) diet, first published in 2015, attempted to refine the MeDi in order to minimize cognitive decline^7^. Though the majority of food groups are similar or identical to those found in the MeDi, the MIND diet uniquely rewards leafy green, berry, and poultry intake while minimizing the consumption of fried food and sweets. Milk, potato, and fruit intake are also discarded. The MIND diet has been associated with up to a 54% reduction in AD incidence^7^ and consistently proves to be more beneficial for cognitive health than the MeDi^8,9^. Despite this success, little research has investigated the effect of the MIND diet on other neurodegenerative diseases. Agarwal et. al (2018) previously showed that higher MIND dietary adherence correlated with reduced incidence and progression of Parkinsonian symptoms during aging^10^, but to date no studies have investigated the potential impact of the diet on patients formally diagnosed with PD. This cross-sectional study examines the relationship between MIND diet adherence and the age of PD onset in a Canadian cohort, and compares the performance of the MIND diet to both MeDi scoring methods.

## METHODS

### Study population and Participant Recruitment

225 participants diagnosed with PD within the last 12 years and 156 control participants were recruited through the Pacific Parkinson’s Research Centre (PPRC) at the University of British Columbia (UBC), Canada, using inclusion/exclusion criteria described previously^11^. Incomplete dietary surveys (n=93) were not included in the analysis, as well as PD participants with no recorded age of onset (n=2), leaving a total of 167 and 119 PD and control participants, respectively. 31 spousal pairs, all of which consisted of one PD and one control participant, were identified from the remaining cohort and excluded from all analyses that involved the control group. The study was approved by the UBC Clinical Research Ethics Board and written informed consent was obtained from each participant.

### Data Collection

All data were self-reported and collected either during a study visit or through an online data collection portal. Age of onset was defined as the age at which the participant first started to experience motor symptoms as recorded in the chart and supported by self-report. Dietary patterns over the past year were assessed using the EPIC-Norfolk Food Frequency Questionnaire (FFQ)^12^, and exercise habits were assessed using the Physical Activity Scale for the Elderly (PASE)^13^. Total energy intake was calculated using FETA^14^ and is reported in kilocalories (kcal). Smoking habits were categorized as current, previous, and never, and blood pressure was self-reported as low, normal, or high. History of diabetes (including gestational diabetes) and cardiovascular disease (CVD) were recorded as true or false, as was family history of PD (confirmed cases in first degree relatives). PASE and PD family history data were only collected from a subset of the PD cohort (n=121 & 123 respectively) as it was included after the study had commenced.

### Diet Scoring

A list of all food groups and the consumption frequencies used for scoring can be found in the Supplementary data (**Tables S1-3**). For all diets, food items that did not fall in any of the listed food groups were discarded. MIND dietary adherence was calculated using the number of servings per food group outlined by Morris et. al^15^, giving MIND scores out of 15 for each participant.

For the OMeDi scoring, food groups were binned as specified by Trichopoulou et. al^5^ and the ratio of monounsaturated to saturated fat intake was calculated using FETA^14^. Participants who consumed below the sex-specific median for dairy and meat were given a score of 1 for the category, or 0 for the remaining categories. Ethanol intake (g/day) was estimated by multiplying the relevant FFQ items by the following ethanol contents: wine (15 g/glass), beer (14.4 g/half pint), ports/liqueurs (10 g/glass), and spirits (9.2 g/shot). A score of 1 was assigned for consumption between 5-25 g/day for women and 10-50 g/day for men. Food group scores were then summed to give the OMeDi score out of 9.

For the GMeDi scoring, food groups were scored out of 5 according to Panagiotakos et. al^6^. Ethanol intake (g/day) was quantified as described above and scored out of 5. Accurate quantifications of olive oil intake were not available; instead, 3 points were added to the total score if olive oil was the primary cooking oil used by the participant. All categorical scores were then summed to give the GMeDi score out of 53.

All dietary tertiles were assigned in a manner that optimized PD participant distribution (**Table S4**).

### Statistical Analysis

All analyses were conducted in R. Univariate associations were queried using Kruskal-Wallis tests for binary variables and linear regression for continuous variables. All multivariate associations between age of onset (response variable) and dietary score (explanatory variable) were queried using linear regression, whereas associations between sex and dietary score used logistical regression. Dietary score was treated as a continuous variable or with tertiles represented as ordinal factors. Nonparametric differences in the distribution of metadata across tertiles were assessed using Kruskal-Wallis tests for continuous variables and chi-square analysis for categorical variables.

## RESULTS

### Cohort Statistics

**Tables 1 and 2** summarize the overall and tertile-based descriptive statistics of the PD and control cohorts respectively. Tables for sex-specific subgroups can be found in the Supplementary (**Tables S5-8**), along with interaction plots of dietary tertiles with each variable (**Figure S1**). Dietary score ranges can be found in **Table S9**.

**Table 1.** PD cohort characteristics.

|  |  | **MIND (/15)** |  |  |  | **Original MeDi (/9)** |  |  |  | **Greek MeDi (/53)** |  |  |  |
| --- | --- | --- | --- | --- | --- | --- | --- | --- | --- | --- | --- | --- | --- |
|  | **All** | **T1** | **T2** | **T3** | **Pval** | **T1** | **T2** | **T3** | **Pval** | **T1** | **T2** | **T3** | **Pval** |
| n (total) | 167 | 49 | 62 | 56 |  | 47 | 77 | 43 |  | 58 | 56 | 53 |  |
| Median Diet Score (IQR) |  | 6 (1) | 7.5 (1) | 9 (1) |  | 2 (1) | 4 (1) | 7 (2) |  | 26 (3) | 31 (2) | 36 (3) |  |
| % Female | 31.7 | 22.4 | 29 | 42.9 | 0.069 | 31.9 | 31.2 | 32.6 | 0.987 | 31 | 32.1 | 32.1 | 0.990 |
| **Age** | 64.9 | 60.9 | 66.5 | 66.6 | **<0.001** | 61.6 | 66 | 66.4 | **0.003** | 62.6 | 64.6 | 67.7 | **0.002** |
| Disease Duration (years) | 6.5 | 6.5 | 7 | 5.8 | 0.146 | 6.2 | 6.5 | 6.7 | 0.677 | 6.4 | 6.8 | 6.1 | 0.408 |
| **Age of Onset** | 58.4 | 54.4 | 59.5 | 60.8 | **<0.001** | 55.4 | 59.6 | 59.7 | **0.023** | 56.2 | 57.8 | 61.6 | **0.001** |
| **Energy Intake (kcal)** | 1659.7 | 1466 | 1748 | 1731 | **0.006** | 1566 | 1605 | 1860 | **0.010** | 1645 | 1632 | 1706 | 0.639 |
| Education (years) | 16.1 | 16 | 16.2 | 16.2 | 0.816 | 15.7 | 16.4 | 16.1 | 0.167 | 16.1 | 15.9 | 16.3 | 0.392 |
| % Smokers (Lifetime) | 40 | 33.3 | 49.2 | 35.7 | 0.177 | 42.6 | 44 | 30.2 | 0.311 | 41.4 | 44.4 | 34 | 0.523 |
| **Exercise Score** | 161.7 | 171.5 | 153.3 | 162.3 | 0.466 | 161.1 | 145.2 | 192.5 | **0.005** | 146.9 | 182.7 | 159.4 | 0.685 |
| % Normal Blood Pressure | 65 | 63.6 | 64.2 | 67.4 | 0.921 | 52.5 | 66.2 | 76.3 | 0.465 | 68 | 57.4 | 69.6 | 0.457 |
| **BMI** | 26.5 | 28.1 | 26.4 | 25.3 | **0.004** | 27.8 | 26.7 | 24.9 | **0.006** | 27.2 | 26.5 | 25.8 | 0.145 |
| Height | 172.8 | 173.7 | 173.7 | 170.8 | 0.139 | 173.2 | 172.5 | 172.7 | 0.942 | 173.5 | 173.4 | 171.3 | 0.464 |
| % Diabetes | 6.4 | 2.9 | 9.8 | 5.9 | 0.482 | 4.2 | 11.1 | 0 | 0.116 | 7.9 | 2.9 | 8.1 | 0.607 |
| **% CVD** | 25.7 | 26.5 | 30.6 | 19.6 | 0.390 | 38.3 | 23.4 | 16.3 | **0.047** | 32.8 | 26.8 | 17 | 0.161 |

The All column represents mean values for the overall cohort, whereas columns T1, T2 and T3 are dietary tertiles. Differences between tertiles were calculated using nonparametric (numerical data) and chi-square tests (categorical data).

**Table 2.** Control cohort characteristics.

|  |  | MIND (/15) |  |  |  | Original MeDi (/9) |  |  |  | Greek MeDi (/53) |  |  |  |
| --- | --- | --- | --- | --- | --- | --- | --- | --- | --- | --- | --- | --- | --- |
|  | All | T1 | T2 | T3 | Pval | T1 | T2 | T3 | Pval | T1 | T2 | T3 | Pval |
| n (total) | 84 | 20 | 30 | 34 |  | 26 | 31 | 27 |  | 20 | 38 | 26 |  |
| Median Diet Score (IQR) |  | 6 (1) | 7.5 (1) | 9 (1) |  | 2 (1) | 5 (1) | 6 (1) |  | 26 (4) | 31 (2) | 37 (4) |  |
| % Female | 60.7 | 35 | 53.3 | 82.4 | **0.002** | 53.8 | 64.5 | 63 | 0.684 | 50 | 68.4 | 57.7 | 0.366 |
| Age | 61.8 | 62.4 | 60.1 | 63.1 | 0.567 | 62.6 | 61.1 | 62 | 0.951 | 66.1 | 57.8 | 64.5 | **0.003** |
| Energy Intake (kcal) | 1569.9 | 1381 | 1603 | 1651 | 0.092 | 1372 | 1650 | 1668 | 0.071 | 1520 | 1597 | 1569 | 0.943 |
| Education (years) | 17.1 | 17.1 | 16.7 | 17.4 | 0.853 | 16.8 | 16.6 | 18.1 | 0.296 | 17.6 | 16.6 | 17.4 | 0.432 |
| % Smokers (Lifetime) | 39.8 | 47.4 | 36.7 | 38.2 | 0.736 | 30.8 | 46.7 | 40.7 | 0.476 | 42.1 | 52.6 | 19.2 | **0.027** |
| Exercise Score | 175.2 | 186.2 | 141.2 | 225.8 | 0.129 | 158.2 | 223.4 | 167.4 | 0.862 | 153.8 | 168 | 192 | 0.645 |
| % Normal Blood Pressure | 60.4 | 50 | 71.4 | 52.9 | 0.440 | 84.6 | 50 | 52.9 | 0.382 | 57.1 | 64 | 56.2 | 0.905 |
| BMI | 26.6 | 28.4 | 25.2 | 27 | 0.254 | 26.8 | 27.5 | 25.6 | 0.582 | 28.2 | 26.1 | 26.2 | 0.209 |
| % Diabetes | 9.1 | 18.2 | 4.3 | 9.5 | 0.421 | 0 | 14.3 | 10 | 0.349 | 12.5 | 10.7 | 5.3 | 0.764 |
| % CVD | 23.8 | 30 | 23.3 | 20.6 | 0.733 | 19.2 | 25.8 | 25.9 | 0.804 | 10 | 28.9 | 26.9 | 0.247 |

The All column represents mean values for the overall cohort, whereas columns T1, T2 and T3 are dietary tertiles. Differences between tertiles were calculated using nonparametric (numerical data) and chi-square tests (categorical data).

PD participants were primarily male (68.3%), were an average of 64.9 years old (SD=8.0), and had begun to experience motor symptoms (referred to as age of onset) an average of 6.5 years previously (SD=3.1). Control participants were only 39.3% male and were slightly younger (M=61.8 years, SD=9.9). PD participants who were older and had later ages of onset had higher adherence to all diets; these correlations remained significant only in the MeDi variants for men and the MIND diet for women. In contrast, age was not significantly associated with any dietary score in the corresponding control groups with the exception of the GMeDi, which was nonlinearly associated, driven by women and likely spurious (Wilcox, p=0.003). High OMeDi adherence correlated with lower CVD incidence and higher exercise scores in the sex-combined PD cohort, while high adherence to both the MIND diet and the OMeDi correlated with higher exercise scores in PD women. High MIND diet adherence also corresponded to higher exercise scores in female controls. PD male adherence to all diets correlated with lower BMIs, though OMeDi and MIND diets were also associated with higher kcal consumption in PD men. High GMeDi adherence corresponded to lower smoking rates in the controls overall and lower CVD rates in male controls.

Women scored 1.1 points higher on the MIND diet than men on average (Wilcox, p<0.001), even after controlling for disease status, kcal, age, and disease duration (logistic regression, p<0.001). Female PD participants appeared to have slightly lower median MIND scores than their control counterparts and vice versa in the male cohort (**Figure S2**); however, these differences were not significant. No other significant associations were observed between other diet scores and sex/disease status.

### MIND Diet Adherence Correlates with Later Disease Onset, Especially Among Women

To facilitate the comparison of model estimates, all dietary scoring systems were adjusted to a 0-10 scale (see **Table S9** for score ranges). Three linear regression models were used to query the relationship between dietary adherence and age of onset: Basic (n=167: disease duration, kcal, sex), Lifestyle (n=121: Basic + smoking, years of education, exercise), and Health (n=123: Basic + high/low blood pressure, diabetes and CVD history, BMI, family PD history). Diet scores were regressed as both continuous adjusted scores and tertiles and the score estimates (β) compared using effect plots (**Figure 2**). Statistics on all models and corresponding regression plots are included in the Supplementary (**Table S10 & Figure S3**).

**Figure 1.**
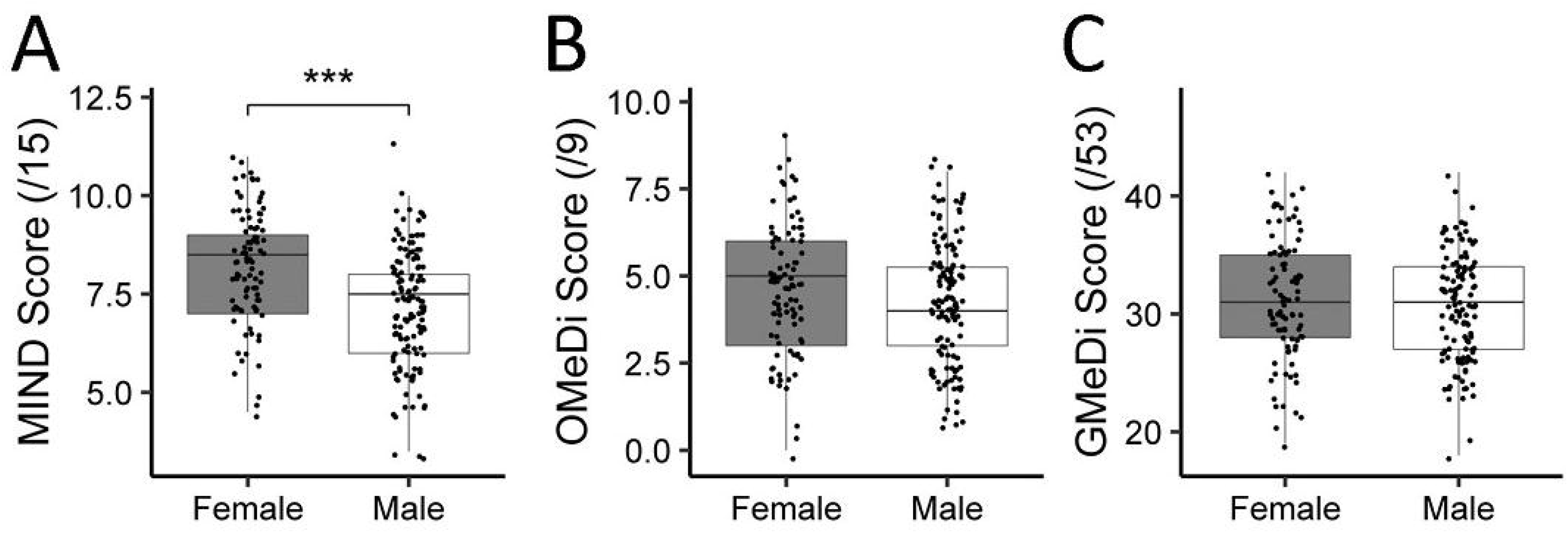
Sex-stratified scores for (A) MIND diet, (B) OMeDi, and (C) GMeDi. *** denotes a p value below 0.001. Figures include both PD and control participants.

**Figure 2.**
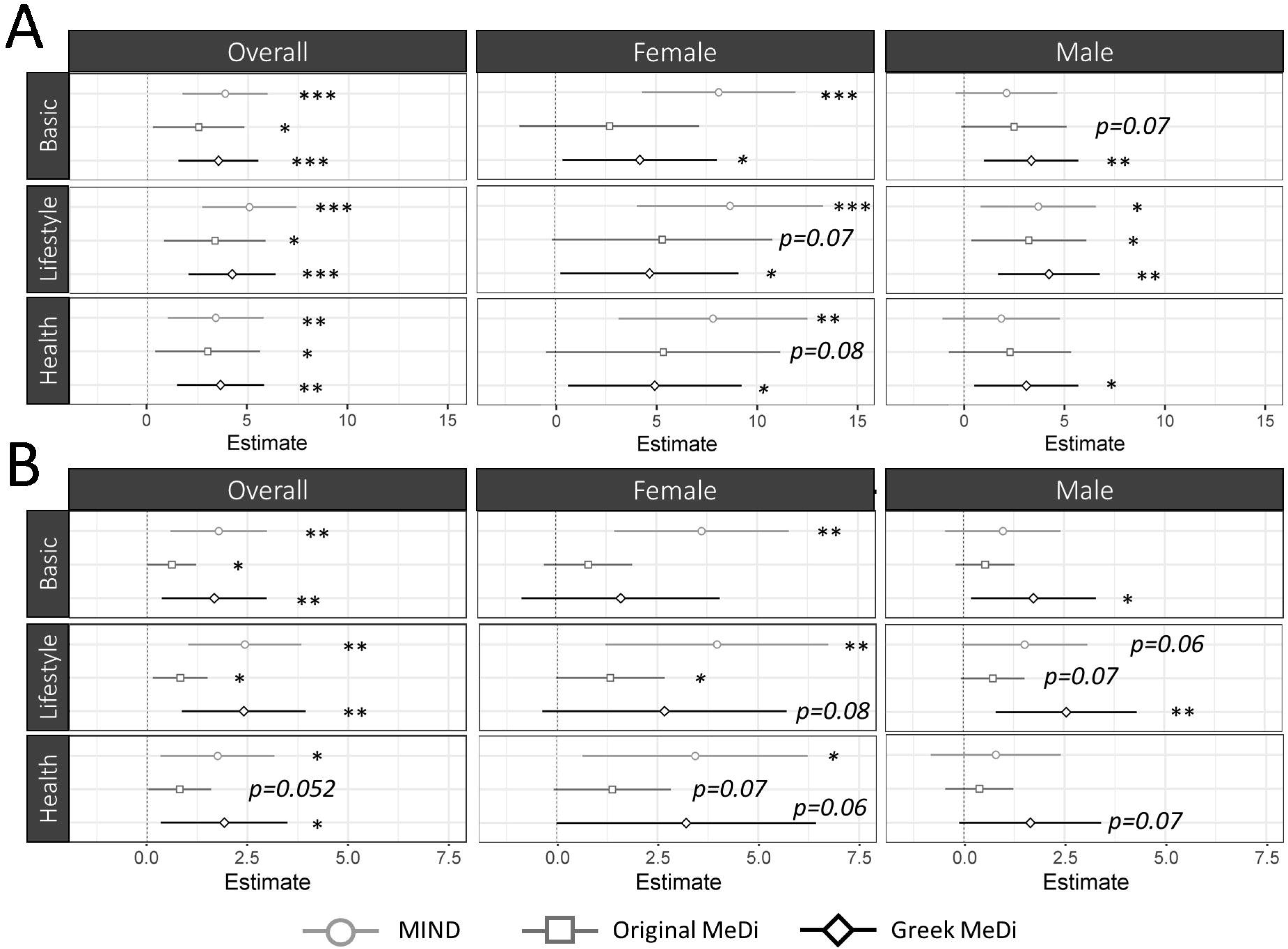
Estimated effects and 95% confidence intervals of the MIND diet, OMeDi, and GMeDi on age of PD onset using (A) fertile and (B) continuous scores. *, ** and *** denote p values below 0.05, 0.01, and 0.001 respectively.

All results discussed compare the estimated difference in age of onset between the lowest and highest dietary tertiles unless otherwise specified (E=2*β, presented as the range of model estimates). While MIND diet adherence correlated most strongly with age of onset in the overall cohort, striking sex-specific effects were revealed upon stratification. Higher MIND diet adherence correlated far more robustly with later onset in women (E=15.6-17.4, p≤0.003) than in men (E=3.6-7.4, p=0.21-0.01) or any other diet in either subgroup (E=4.6-10.8, p<0.25). The GMeDi model in the female subgroup also reached significance, though to a lesser degree (E=8.4-9.8, p=0.05-0.03). In men, the GMeDi correlated most consistently with age of onset (E=6.2-8.4, p=0.02-0.002) and was the only diet to remain significantly associated across every model. The MIND diet was only weakly correlated with age of onset (E=3.6-7.4, p=0.21-0.01), performing similarly to the OMeDi (E=4.6-6.4, p=0.15-0.03). Similar trends were observed between the tertile and continuous datasets, though OMeDi effect sizes were far smaller due to their wider score range (**Table S9**).

## DISCUSSION

In this cross-sectional study, higher adherence to the MIND diet was significantly associated with a higher age at disease onset, especially in women, with a difference of up to 17.4 years between the highest and lowest tertiles of diet adherence. In men, the GMeDi was consistently more significant than the MIND diet and the OMeDi across models; still, no diet was associated with more than a 7.1-year average difference in age of onset between low and high tertiles in men. While female participants experienced only slightly larger MeDi effect sizes compared to male participants, the average effect size of the MIND diet in females was over 3 times that of the males and surpassed all MeDi effect sizes, suggesting that its dietary components are better suited to possibly delaying PD onset than MeDi in a female-specific manner.

Similarly, only the MIND diet showed any interaction between sex and diet score, despite the fact that neither the MIND diet nor the GMeDi normalizes food intake by sex. Female participants adhered significantly closer to the MIND diet than males, even after correcting for age, disease status and duration, and kilocalorie consumption, indicating that the higher MIND score is not simply due to differences in food volume. As the sex difference was similar between the PD and control groups (β=1.0 & 1.2 respectively), it is unlikely that this effect is an artefact of any sex-specific dietary shifts that may occur upon PD diagnosis. This tendency for females to adhere more strongly to the MIND diet may contribute to their lower rate of PD incidence.

An analysis of two large US cohorts found that while GMeDi adherence was only weakly associated with reduced PD risk (p=0.07), the ‘prudent’ dietary pattern was slightly more strongly associated (p=0.04)^16^. Interestingly, this prudent pattern promoted several items such as poultry and leafy vegetables in a manner similar to the MIND diet rather than either MeDi.

Several other studies have also found negative correlations between PD status or risk and adherence to MeDi- or MIND-type diets^17,18^. These results are at odds with the present findings, which found no significant interactions between diet and disease status. It is possible that any dietary differences that may have existed between PD and control participants prior to disease onset are corrected upon disease diagnosis in a sex-independent manner; however, the strength of the interactions between age of onset and dietary scores suggest that any dietary shifts that may occur upon diagnosis do not significantly affect the results.

Apart from age and kcal consumption, the only sex-specific associations noted between PD dietary scores and the model covariables involved exercise in women and BMI in men; thus, the corresponding Lifestyle and Health models were presumed to be the most accurate predictors of dietary effects in women and men respectively. Though all three models (Basic, Lifestyle, Health) produced similar diet rankings, the Health model resulted in slightly lower average effect sizes compared to the Lifestyle model in men. It is well documented that the MeDi imparts significant cardiovascular benefits^1,19^, some of which are sex-dependent; for example, improved insulin homeostasis has been observed only in men^19^. Indeed, significantly reduced CVD incidence was noted in those with high OMeDi scores (**Table 1**), and trended similarly for the majority of other diet/sex combinations. If MeDi-type diets delay PD onset in part via their beneficial cardiovascular effects, then controlling for CVD may reduce the apparent effect of the diets, especially in men. Similarly, the higher and more statistically significant effect sizes observed in the Lifestyle model in men support the notion that the model covariables are significant disease-modifying elements. While smoking has long been associated with reduced PD incidence^20^, the impact of exercise has not been fully explored. A growing number of studies, including the large-scale FINGER study^21^, have suggested that exercise may be an effective way to reduce neurological decline, especially as part of a combinatorial therapeutic approach^22^.

Although adherence to all diets was strongly associated with lower BMIs in male PD participants, it was also positively associated with higher kilocalorie consumption with no significant changes in exercise habits. As the majority of food groups in each diet reward increased consumption, it is possible that taller people naturally score higher than shorter people while still maintaining similar or lower BMIs due to their higher energy requirements. However, no correlations were found between diet scores and height (**Table 1**). It is likely that people with low dietary scores consume more foods that are not captured by the FFQ, such as prepackaged meals, and thus their kcal consumption is underestimated.

To the best of our knowledge, this is the first study to examine the role of the MIND diet in a strictly PD cohort. Our female PD-specific findings mirror previous research in AD and cognitive decline, where the MIND diet has repeatedly proven more effective than MeDi as a preventative measure over several different mixed-sex study cohorts^7,9,15^. Interestingly, women represent two thirds of all AD cases and may experience more severe cognitive deficits than their male counterparts^23^. The observed effects of the MIND diet in AD and in women with PD suggest that the diseases share similar sex-dependent mechanisms which may be modulated by dietary intake. Several previous studies have indicated that certain effects of MeDi are sex-specific in neurotypical cohorts, such as inflammation^24^ and reduced CVD risk^19^ as mentioned previously.

In contrast, few studies have previously identified sex-based differences related to the MIND diet^25^. Future work will investigate the effects of the MIND diet on other elements of PD etiology including disease progression, inflammatory markers, and gastrointestinal symptoms such as constipation and dysbiosis.

These findings also corroborate a recent longitudinal study by Agarwal et. al^10^, where participants in the highest MIND dietary tertile developed parkinsonism at a rate 42% below that of the lowest tertile over an average observation period of 4.6 years. This analysis studied the RUSH Memory and Aging Project (MAP) cohort, which was also used to identify a positive correlation between MIND diet adherence and reduced incidence/progression of cognitive decline^15^ and AD^7^. Importantly, the MAP cohort is 75% female. Though no sex-specific effects were reported in these studies, the high proportion of women suggests that the results are more reflective of female physiology. Beyond the sex ratio, the lack of sex-specific effects observed may be due to several factors: firstly, the advanced age of the MAP participants (approximately 80, 15 years older than the present cohort) suggest that the sex specificity observed here may be particularly relevant for the delay of neurodegenerative disease in early/mid senium. Additionally, parkinsonism is an umbrella term that does not constitute a diagnosis of PD. In the corresponding study, 43% of the participants developed parkinsonism over a mean follow up of 4.6 years, which is an order of magnitude higher than the 10-year PD risk estimate for males aged 75 (2.6%)^26^. It is possible that the more inclusive definition of parkinsonism in the analysis masked any sex-specific effects that may be particular to PD. Finally, the methods used to detect sex-specific effects were not specified, and so direct comparisons cannot be made between studies.

Due to the complexity of the diets, the key elements that drive their beneficial effects are poorly understood. It is believed that the power of the diets is due to a complex range of metabolites acting upon multiple disease elements; however, significant progress has been made to identify key molecules and metabolites that act upon neurodegenerative diseases in reproducible ways. Leafy greens and berries, which are specific to the MIND diet, are rich in antioxidants such as carotenoids, flavonoids, folate, and vitamins C and E, some or all of which have been associated with lower PD/parkinsonism risk and reduced disease progression in both animal models and human cohorts^27-31^. Conversely, the MeDi diets restrict the intake of all dairy, while the MIND diet penalizes only cheese and butter/margarine consumption. Milk consumption has been repeatedly identified as a risk factor for PD, possibly due to increased pesticide exposure; its omission from the MIND diet may contribute to the reduced efficacy of the diet observed in the male cohort. Overall, determining the subtle differences in the metabolic profiles of the different diets may help to unravel elements of PD etiology that are modified by diet in a sex-specific manner.

Several limitations should be noted with this study. Firstly, all dietary data are cross-sectional, where only one FFQ was analyzed per participant; in addition, the analysis assumes that the dietary habits of each participant have not significantly changed over their lifetime. Though there were no differences found between PD and control dietary scores, a prospective study would be required in order to ensure that all disease-related dietary fluctuations are accounted for. Secondly, the berry food group included in the MIND diet is underrepresented by the FFQ, as the only related question assesses strawberries, raspberries, and kiwi fruits and disregards other common berries such as blueberries. Lastly, there is a strong correlation between the age of the participant and age of onset (p<0.001) (**Figure S4**), meaning that any interactions between age and dietary score are misattributed to age of onset. This strong interaction is a result of the study design: only patients who had been diagnosed with PD for 12 years or less were included, resulting in a narrow disease duration range (M=6.5 years, SD=3.1). Despite this limitation, no significant linear correlations were found between age and diet scores in the controls (**Table S11**) and the results presented here are thus believed to be valid.

We have captured a strong, female-driven correlation between MIND diet adherence and delayed PD onset in a manner similar or superior to the MeDi. The sex specificities presented here are novel and may prove to be an important contributor to the sex differences observed in PD. Overall, these data paint a compelling rationale for interventional and animal-based studies that investigate the direct impact of the diet on PD etiology in a sex-specific manner. This study should be repeated in a larger, preferably prospective cohort in order to confirm these findings. Future work will investigate the effect of the diet on other PD symptoms including gut microbial dysbiosis, disease progression, constipation, cognition, and other factors.

## Data Availability

No patient data is being released with this manuscript.

## ACKNOWLEDGEMENTS

We thank Dr Seti Boroomand, PhD and Faezah Kharazyan from the Borgland Family Brain Tissue and DNA Bank, Biobank at the Djavad Mowafaghian Centre for Brain Health, for their invaluable assistance. We would also like to thank all study participants for their contributions to this study, along with all PPRC clinicians and staff members who facilitated the patient recruitment process. We would also like to acknowledge the contribution of the staff and participants of the EPIC-Norfolk Study.

## AUTHORS’ ROLES

1. Research project: A. Conception, B. Organization, C. Execution;
2. Statistical Analysis: A. Design, B. Execution, C. Review and Critique;
3. Manuscript: A. Writing of the first draft, B. Review and Critique.

A.M.R.: 1A-C, 2A-C, 3A-B.

A. C.Y., E.G.: 1B-C, 3B.

K.S.: 1A-C.

M.S.C.: 2A,C, 3B.

D.K., L.H.F., M.M.: 1B-C.

B.B.F.: 1A-C, 2A,C, 3B

S.A.C.: 1A-C, 2A,C, 3B

